# Radical Pericardiectomy and Use of Cardiopulmonary Bypass for Constrictive Pericarditis

**DOI:** 10.1101/2024.06.04.24308462

**Authors:** Marijan Koprivanac, Karolis Bauza, Nicholas Smedira, Gosta Pettersson, Shinya Unai, Paola Barrios, Nicholas Oh, Filip Stembal, Valentina Lara-Erazo, Edward G. Soltesz, Faisal G. Bakaeen, Haytham Elgharably, Milind Desai, Tom K. Wang, Penny L Houghtaling, Lars G. Svensson, Marc Gillinov, Kenneth McCurry, Douglas R. Johnston, Eugene H. Blackstone, Allan Klein, Michael Z. Tong

**Author notes:** **Address for correspondence:** Marijan Koprivanac, MD, The Center for Diagnosis and Treatment of Pericardial Diseases, Miller Family Heart, Vascular & Thoracic Institute, Cleveland Clinic, 9500 Euclid Ave., Desk J4-477, Cleveland, OH 44195. Co-directors, The Center for Diagnosis and Treatment of Pericardial Diseases, Miller Family Heart, Vascular & Thoracic Institute, Cleveland Clinic (Marijan Koprivanac, Allan Klein).

## Abstract

**Background:** Pericardiectomy is definitive treatment for constrictive pericarditis. However, extent of resection (radical versus partial) and use of cardiopulmonary bypass (CPB) are debated.

**Objectives:** To determine the association of extent of pericardial resection and use of CPB with outcomes.

**Methods:** From January 2000 to January 2022, 565 patients with constrictive pericarditis underwent radical (n=445, 314 [71%] on CPB) or partial (n=120, 67 [56%] on CPB) pericardiectomy at Cleveland Clinic. Outcomes stratified by extent of pericardial resection and use of CPB were compared after propensity-score matching.

**Results:** Both radical pericardiectomy and CPB use (67% [381/565]) increased over time. Among 88 propensity-matched pairs (73% of possible matches), immediate postoperative cardiac index increased (*P*<0.001) in both groups by a median of 1.0 L•min-1•m-2. There were no significant differences between radical versus partial resection groups in occurrence of reoperation for bleeding (2.3%, [2/88] vs. 0, *P*=.50). Median postoperative hospital length of stay was 10 versus 8.5 days (*P*=.02). Operative mortality was 9.1% (8/88) versus 6.8% (6/88) (*P*=.58). 10-year survival was 54% versus 41%, with a higher propensity-adjusted hazard ratio after partial resection (1.9, 95% CI 1.2-3.1).

**Conclusions:** When surgical intervention is deemed necessary, radical — rather than partial — resection for constrictive pericarditis can be performed with low surgical mortality and morbidity. Radical pericardiectomy can be accomplished on CPB and results in better long-term survival.

**CLINICAL PERSPECTIVES:**

- Patients with constrictive pericarditis require a multidisciplinary approach involving primarily a cardiologist and cardiac surgeon, and other disciplines like gastroenterology since liver cirrhosis from increased central venous pressure and congestion is common, or immunology for evaluation of possible autoimmune etiology.
- Communication is critical in managing patient expectations after pericardiectomy, especially linking etiology to short- and long-term outcomes in this complex patient population.
- Radical pericardiectomy should be the gold standard for treating patients with constrictive pericarditis.
- Routine use of cardiopulmonary bypass is safe and enables the radical pericardiectomy surgery and should be recommended in the guidelines.

## INTRODUCTION

> ***Constrictive pericarditis is caused by pericardial inflammation, followed by fibrosis, adhesion, and thickening, leading to loss of elasticity.***^1,2^ ***Etiology varies from idiopathic to post-cardiotomy to radiation therapy.***^3^ ***Untreated, it can lead to important morbidity and mortality.***^4,5^ ***(Supplemental Appendix A.1)***

Compared to medical treatment, surgical pericardiectomy yields superior survival, greater functional improvement, and fewer recurrences.^6, 7,8^ However, surgical approach and extent of resection varies, in part from absence of a standardized surgical technique and perception of high surgical risk.^7,9^ This leads to hesitancy to operate on mildly symptomatic patients, and those with advanced disease seem to not benefit from surgery or are deemed too high risk.^1^

European guidelines recommend thorough pericardial resection to remove all sites of inflammation.^10^ They suggest using cardiopulmonary bypass (CPB) only when necessary. However, the association of CPB use with higher mortality is not consistent ^4^, with recent studies reporting no association.^7,11,12,13^

We therefore have evaluated the association of extent of pericardial resection — radical or partial — and use of CPB with early postoperative complications and long-term survival outcomes of patients with constrictive pericarditis who received surgery.

## PATIENTS AND METHODS

From January 2000 to January 2022, 565 patients with constrictive pericarditis underwent pericardiectomy, comprising 445 radical and 120 partial resections, at Cleveland Clinic, Cleveland, Ohio. Patients with recurrent pericarditis, pericardial tamponade or effusion, or tumor invasion into pericardium were excluded. Mean age was 61±12 years (Table 1). Advanced heart failure was present in 40% of those undergoing radical resection and 46% undergoing partial resection. Twelve patients undergoing radical and 10 undergoing partial pericardiectomy had previous pericardiectomy for constrictive pericarditis, and 212 patients (150 radical and 62 partial pericardiectomy) had prior cardiovascular surgery.

**Table 1.**
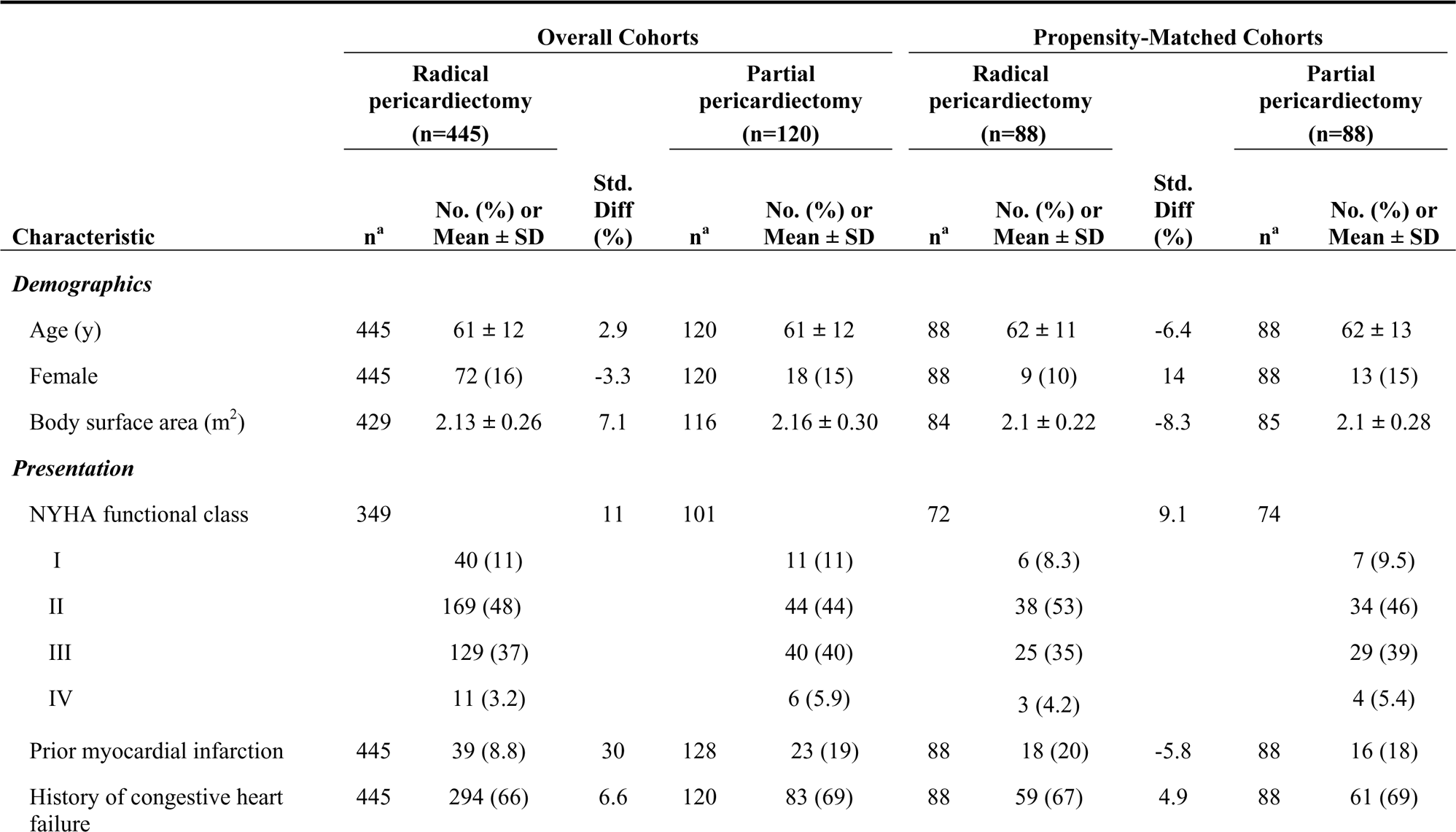
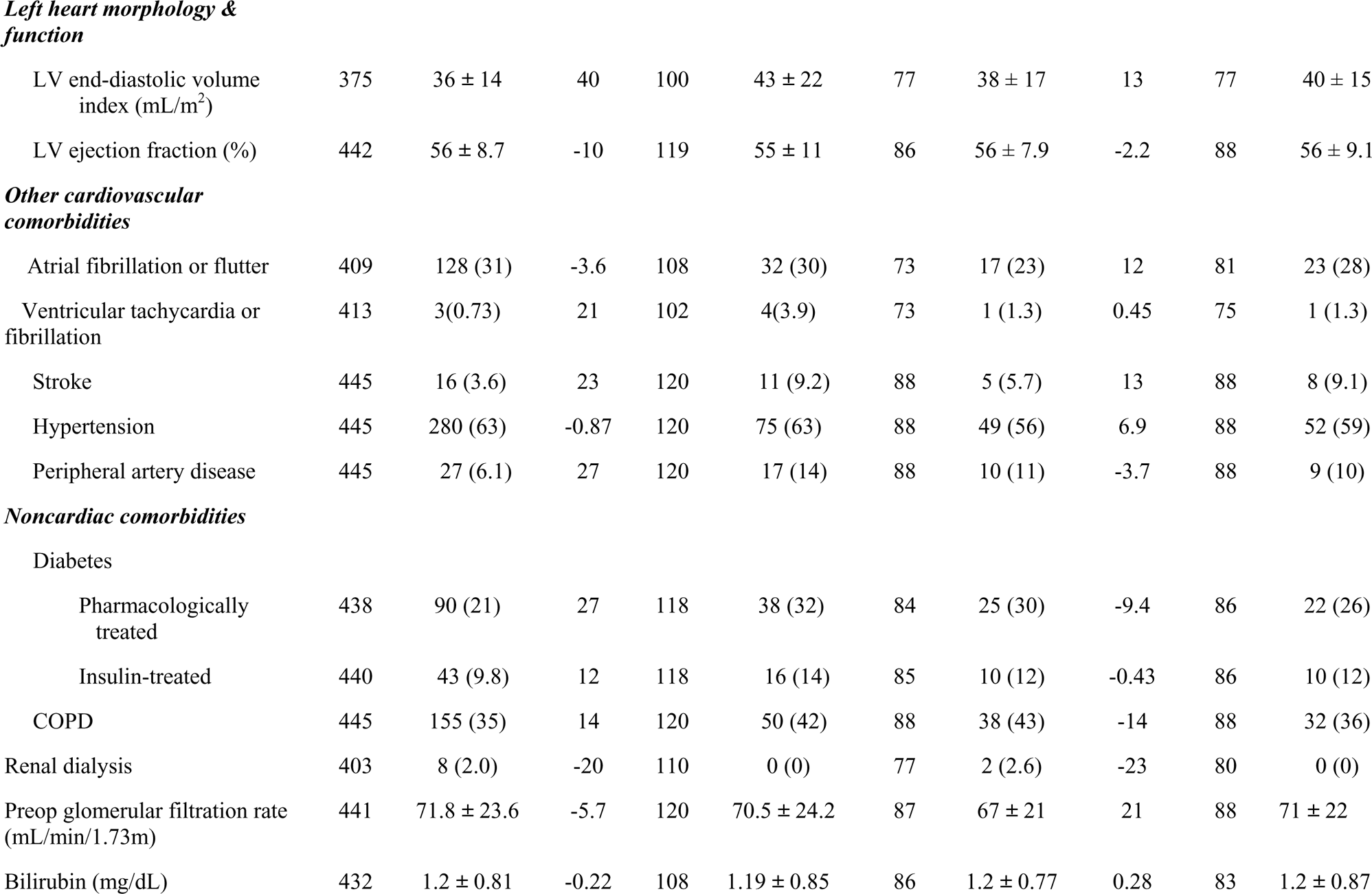
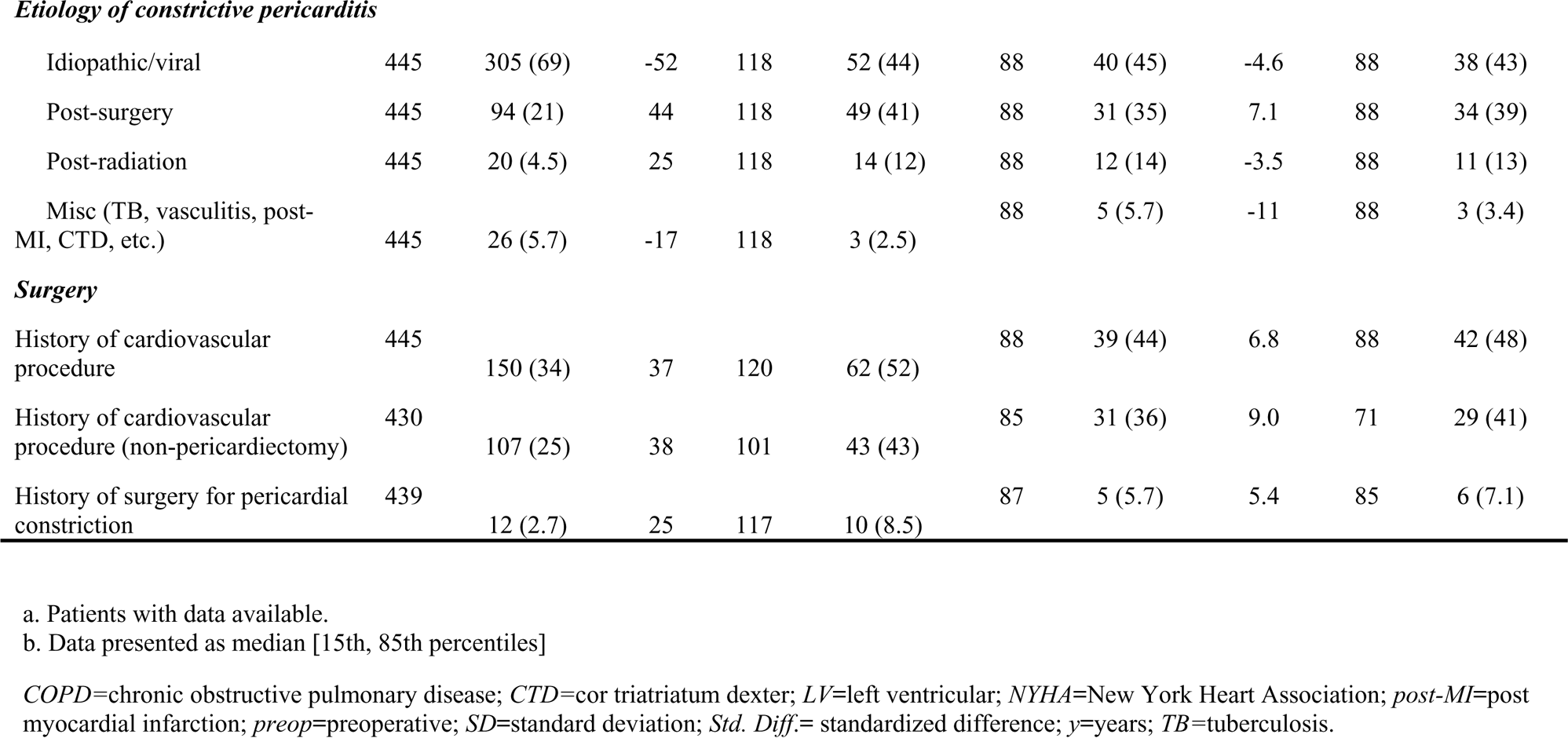
Baseline Characteristics of Patients with Constrictive Pericarditis Undergoing Radical and Partial Pericardiectomy, Original and Propensity-Matched Cohorts.

Etiology of constrictive pericarditis was idiopathic or viral in 63% (n=357), post-cardiotomy in 25% (n=143), radiation in 6% (24), and miscellaneous in 5.1% (n=29) (Table 1). Its diagnosis was established according to American Society of Echocardiography Clinical Recommendations for Multimodality Cardiovascular Imaging of Patients with Pericardial Disease.^14^

### Pericardiectomy

#### Radical Pericardiectomy

We considered radical pericardiectomy as the removal of all pericardium, including a strip along the phrenic nerves, as previously described.^1^

#### Partial Pericardiectomy

We considered partial pericardiectomy as one with less than the entire removal of the pericardium. Partial pericardiectomy included operations that were reported as complete but did not remove all of the pericardium.

### Endpoints

#### Operative Morbidity and Mortality

Postoperative hospital morbidity was defined in accordance with The Society of Thoracic Surgeons National Database. Operative mortality included postoperative in-hospital deaths and deaths after hospital discharge up to 30 days after surgery.

#### Postoperative Cardiac Index

Postoperative cardiac index was measured immediately after surgery in the intensive care unit. Preoperative and postoperative cardiac index data was summarized and compared between radical and partial resection groups. Paired t-test was used to test absolute change (postoperative – preoperative) between groups. Postoperative cardiac index was grouped with the lowest 15th percentile and highest 85th percentile values to stratify survival curves.

#### Long-term Mortality

All-cause, time-related mortality included operative mortality. Survival was estimated nonparametrically by the Kaplan-Meier method and parametrically by a multiphase hazard model.^15^ Vital status was obtained by systematic follow-up at 6 months and 1, 5, and 10 years. Median follow-up was 1.5 years with 25% followed >6.3 years and 10% for >11 years. In matched cohorts, 25% were followed for >7.4 years and 10% for >11 years.

### Data

Clinical data were extracted prospectively for quality assurance. Cleveland Clinic’s Institutional Review Board approved use of these data for research, with patient consent waived (IRB #5001; approved 2/26/2021).

#### Data Analysis

Analyses were performed using SAS statistical software (SAS v9.4, SAS, Inc., Cary, NC) and R software version 4.2.3 (http://www.R-project.org). Continuous variables are summarized as mean ± standard deviation, or as equivalent 15th, 50th (median), and 85th percentiles when data distribution was skewed. Categorical variables are summarized as frequencies and percentages. Asymmetric 68% confidence limits are consistent with ±1 standard error.

#### Propensity-Score Rationale, Development, and Matching

Differences in patient and procedural characteristics of those undergoing radical versus partial pericardiectomy precluded direct comparison of outcomes. Therefore, propensity matching was used to reduce selection bias (Supplemental Figure S1).^16^

##### Managing missing data

Before identifying factors related to extent of resection, we used 5-fold multiple imputation for missing data values using multivariate imputation by chained equations. Variable selection used 1 imputed data set. Regression coefficients for final model variables and their variance–covariance matrix were estimated for each imputed data set and combined using Rubin’s method.^17^

##### Parsimonious model

Multivariable logistic regression was performed to identify factors associated with partial pericardiectomy from those considered in Supplemental Appendix 1. Variable selection used machine learning with automated analysis of 1,000 resampled (bootstrapped) data sets followed by tabulating the frequency of occurrence of variables included in each model at *P*≤.05. We retained factors that occurred in 50% or more of the bootstrap models (Supplemental Table S1) (C=.77).^18^

##### Propensity model

The parsimonious model was augmented with other demographic and comorbidity variables to account for unrecorded factors to form a semi-saturated model of 67 variables (C=.84). Five such models with identical factors were constructed, 1 for each multiple imputation dataset, and a propensity score for each patient was calculated from each and averaged. For propensity matching, the maximum allowable difference between patients for the propensity score was set to 0.2 times the standard deviation (SD) of the logit of the propensity score (0.2×SD [logit(propensity score)])^19^, yielding 88 matched pairs out of 120 possible pairs (73% of partial pericardiectomy cases) (Table 1).

##### Comparison

Wilcoxon signed-rank and chi-square tests were used to compare continuous and categorical in-hospital outcome variables in the matched cohorts. Time-related survival was compared between matched groups and by adjusting for propensity score in the overall cohort.

## RESULTS

### Overall Experience

The proportion of radical pericardiectomies increased over the study period from 60% in 2001 to 95% in 2021 (Figure 1). Hospital mortality for all patients undergoing pericardiectomy decreased from 14% in 2000 to 4% in 2021 (Supplemental Figure S2). The trend in CPB use increased from 37% to 100% of cases from 2000 to 2021 (Figure 2). Unadjusted survival varied according to etiology of constrictive pericarditis, with lowest survival in the post-radiation patient group (Supplemental Figure S3). Low postoperative cardiac index was associated with poor survival (Supplemental Figure S4).

**Figure 1.**
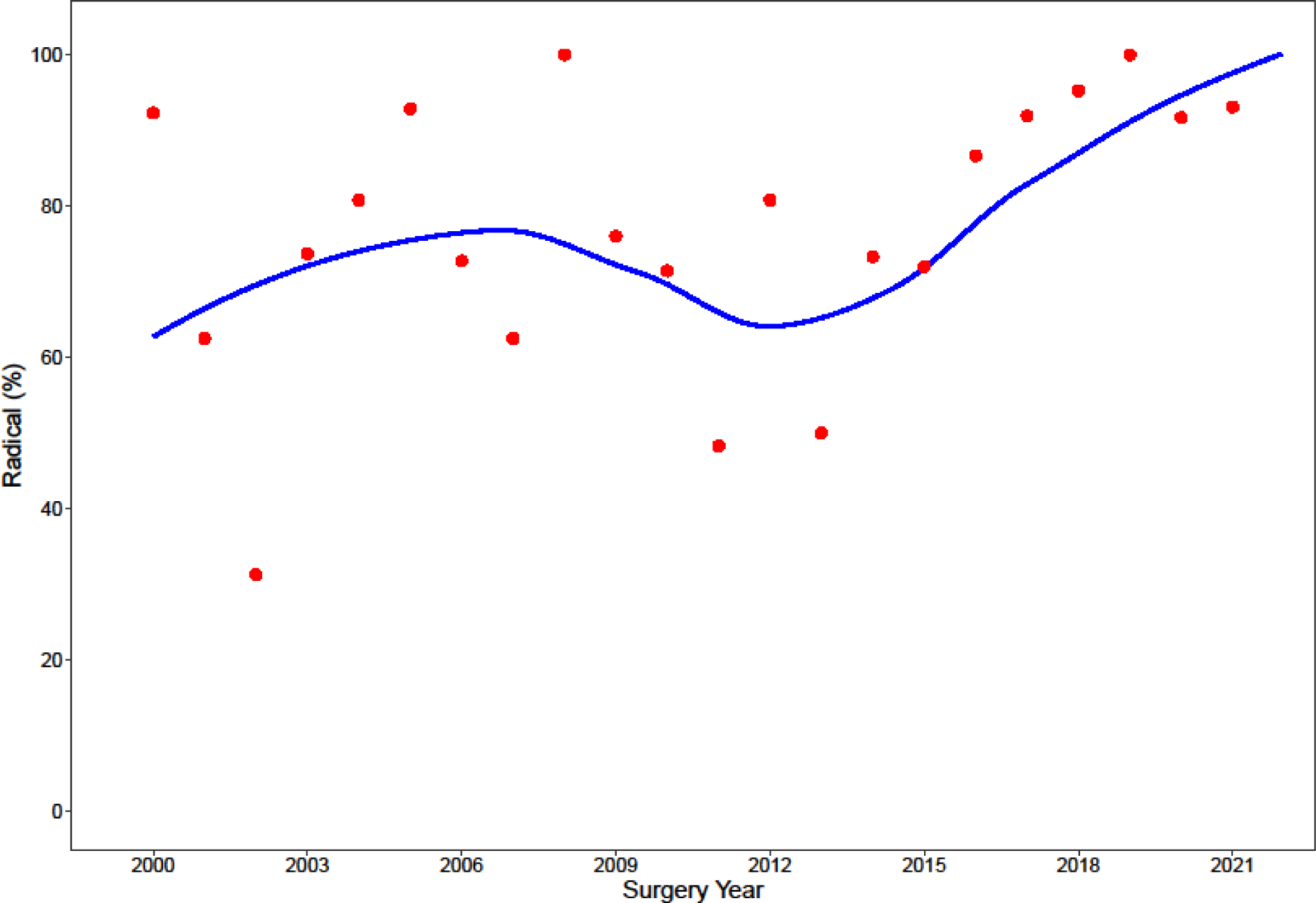
Temporal Trend of Radical Pericardiectomy by Year. Symbols represent yearly percentages, with a smoothing trend line.

**Figure 2.**
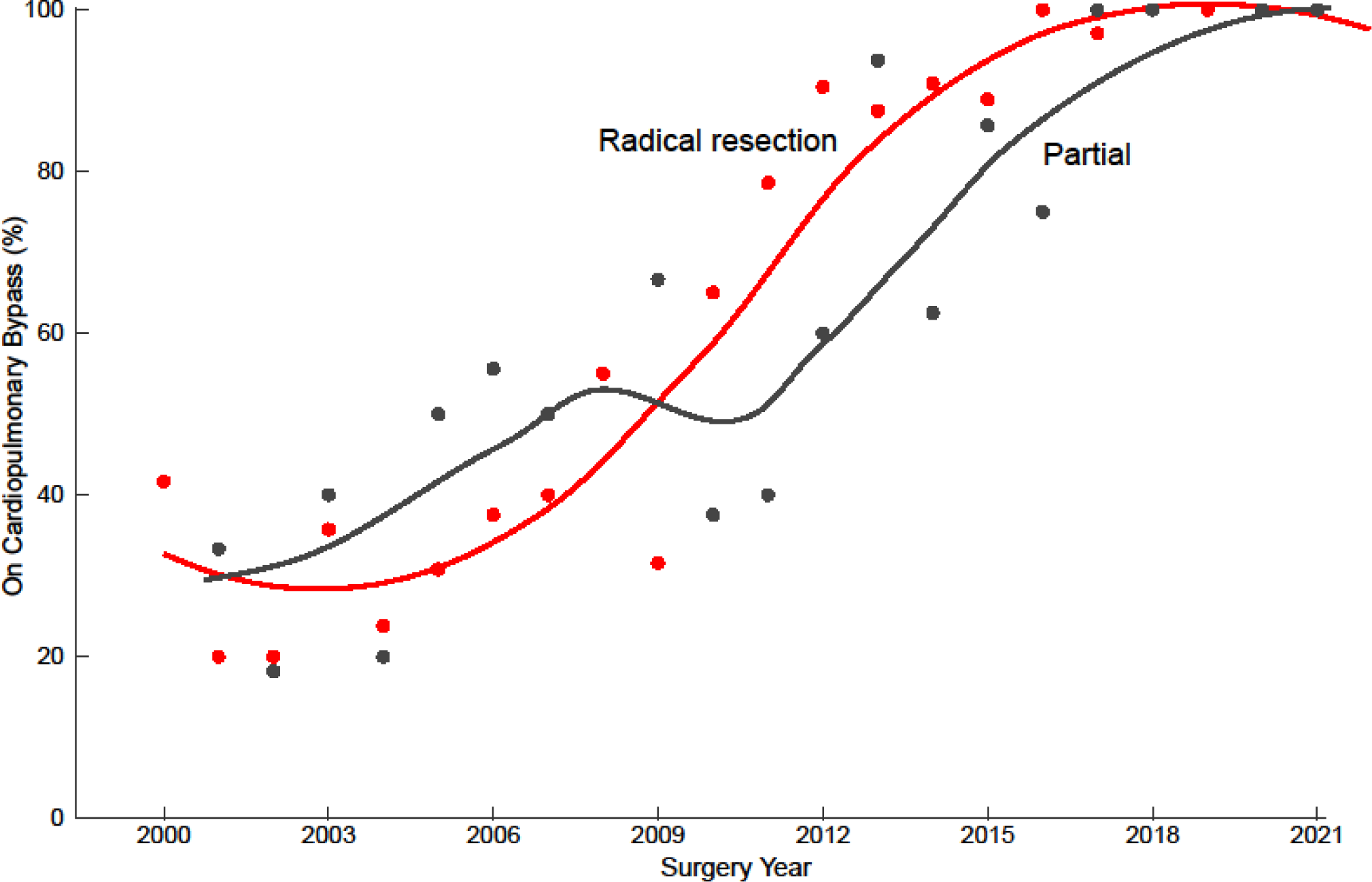
Temporal Trend of Pericardiectomy on Cardiopulmonary Bypass by Year. Symbols represent yearly percentages, with a smoothing trend line. Red-Radical resection; Black-Partial resection.

### Radical Pericardiectomy

Mean age of patients who underwent radical pericardiectomy was 61±12 years. This group comprised 72 (16%) females, 280 (63%) with hypertension, 90 (21%) with pharmacologically treated diabetes, 43 (9.8%) insulin-treated, 155 (35%) with chronic obstructive pulmonary disease (COPD), and 8 (2%) with chronic dialysis (Table 1).

Etiology of constrictive pericarditis was idiopathic or viral in 305 (69%) patients, post-cardiotomy in 94 (21%), post-radiation in 20 (4.5%), and miscellaneous in 26 (5.7%).

Mean left ventricular ejection fraction was 56±8.7%. Mean preoperative cardiac index was 1.86±0.62 L•min^−1^•m^−2^ (Table 2).

**Table 2.**
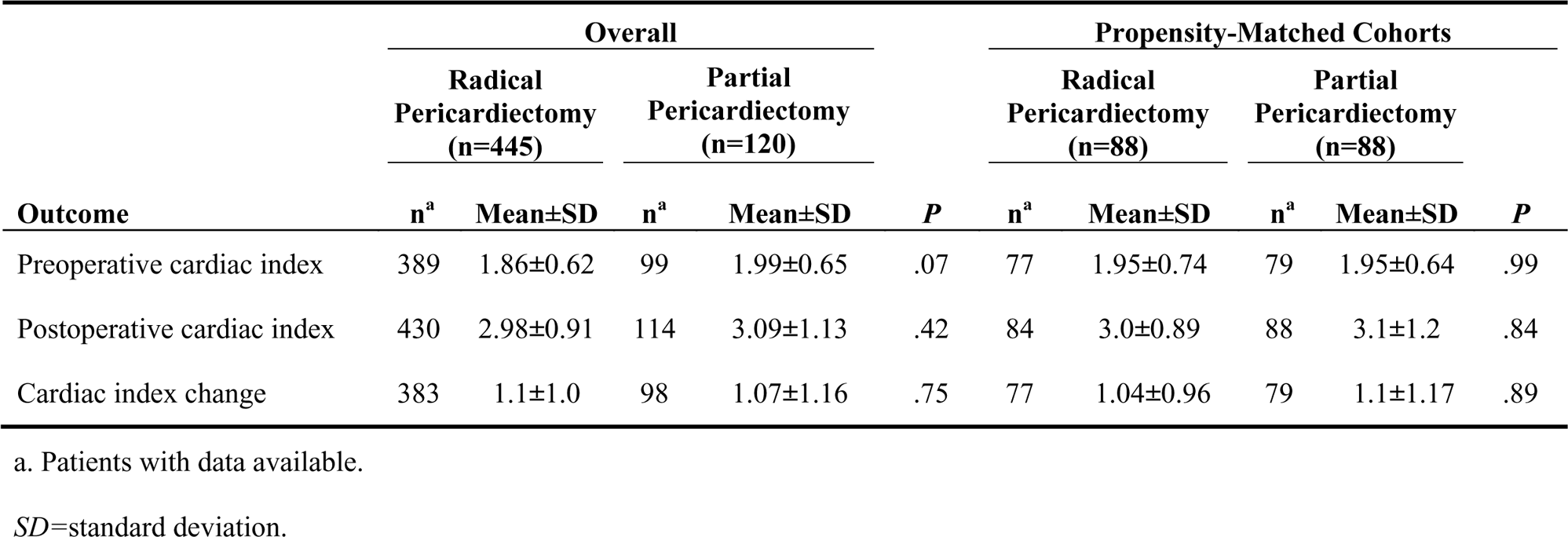
Cardiac Index.

CPB was used in 314 (71%) (Table 1). Intraoperatively, pericardial calcifications were observed in 214 (48%) patients (Table 3). Mean postoperative cardiac index was 2.98±0.91 L•min^−1^•m^−2^.

**Table 3.**
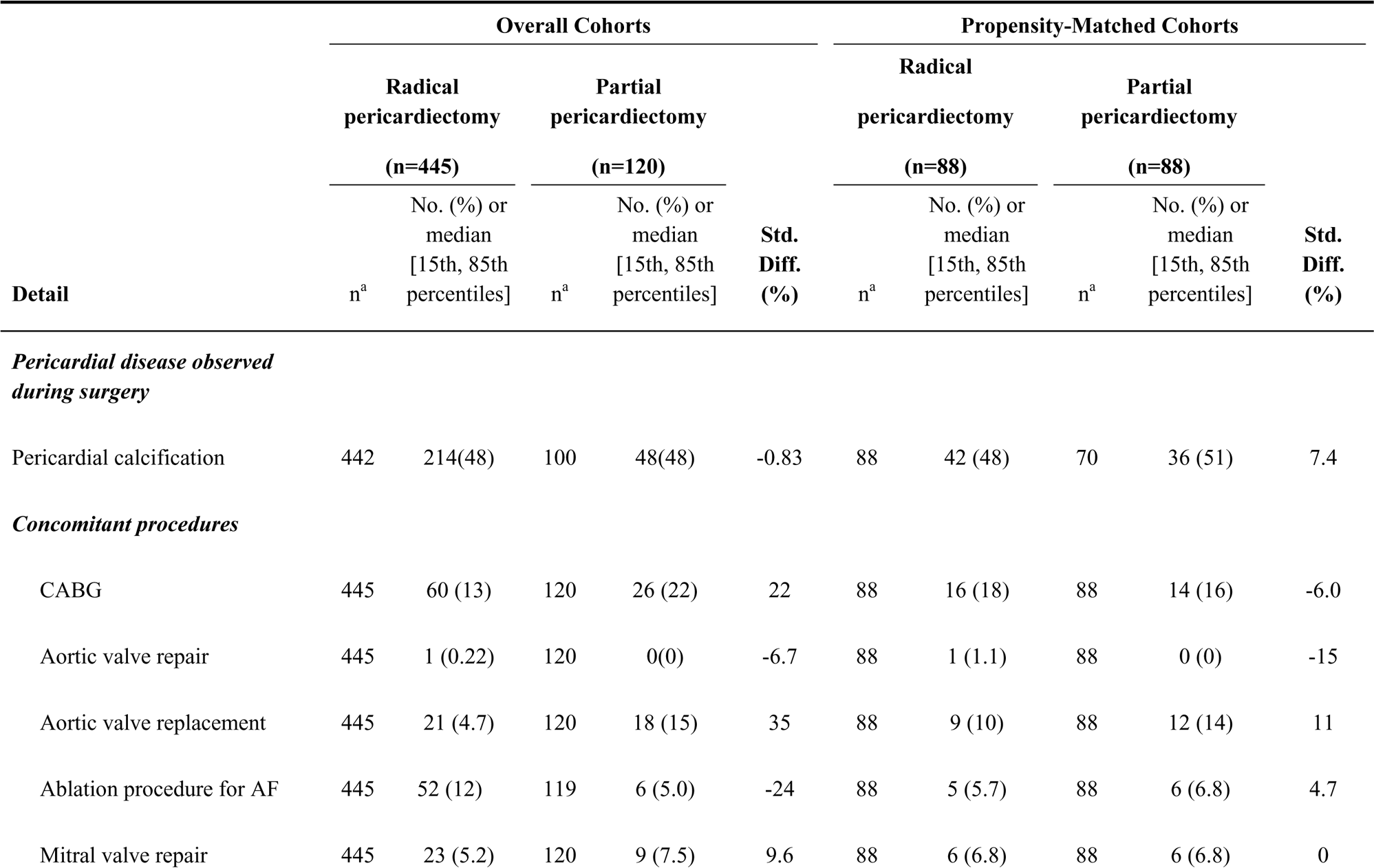
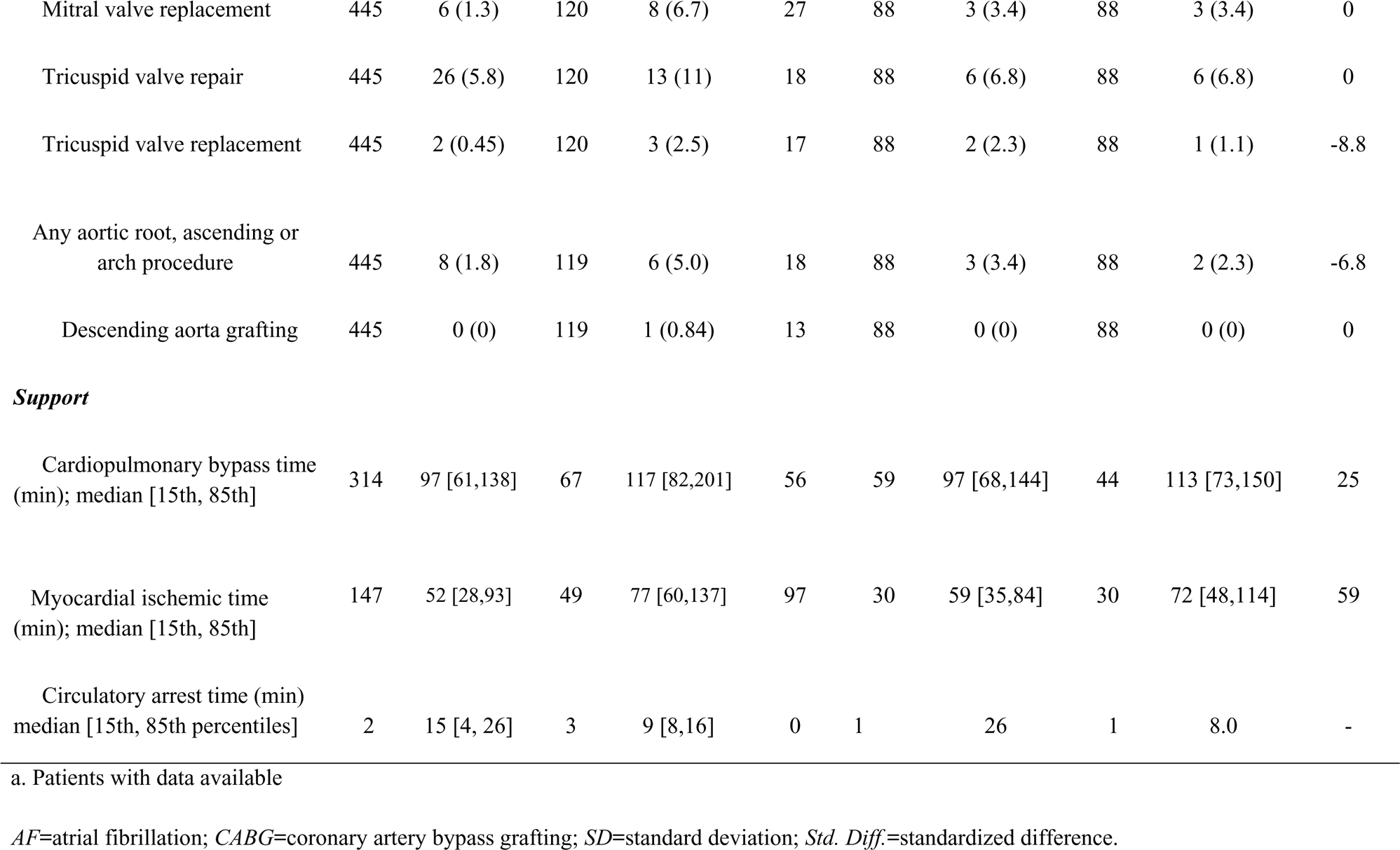
Operative Details.

Following radical pericardiectomy, reoperation for bleeding or tamponade was performed in 0 of 131 of patients whose pericardiectomy was performed without CPB and 13 of 314 (4.1%) in whom CPB was used (Table 4). Intra-aortic balloon pump (IABP) was used for postoperative support in 6 (1.3%). Extracorporeal membrane oxygenation (ECMO) was used in 12 (2.7%) and left ventricular assist device (LVAD) in 3 (0.7%).

**Table 4.**
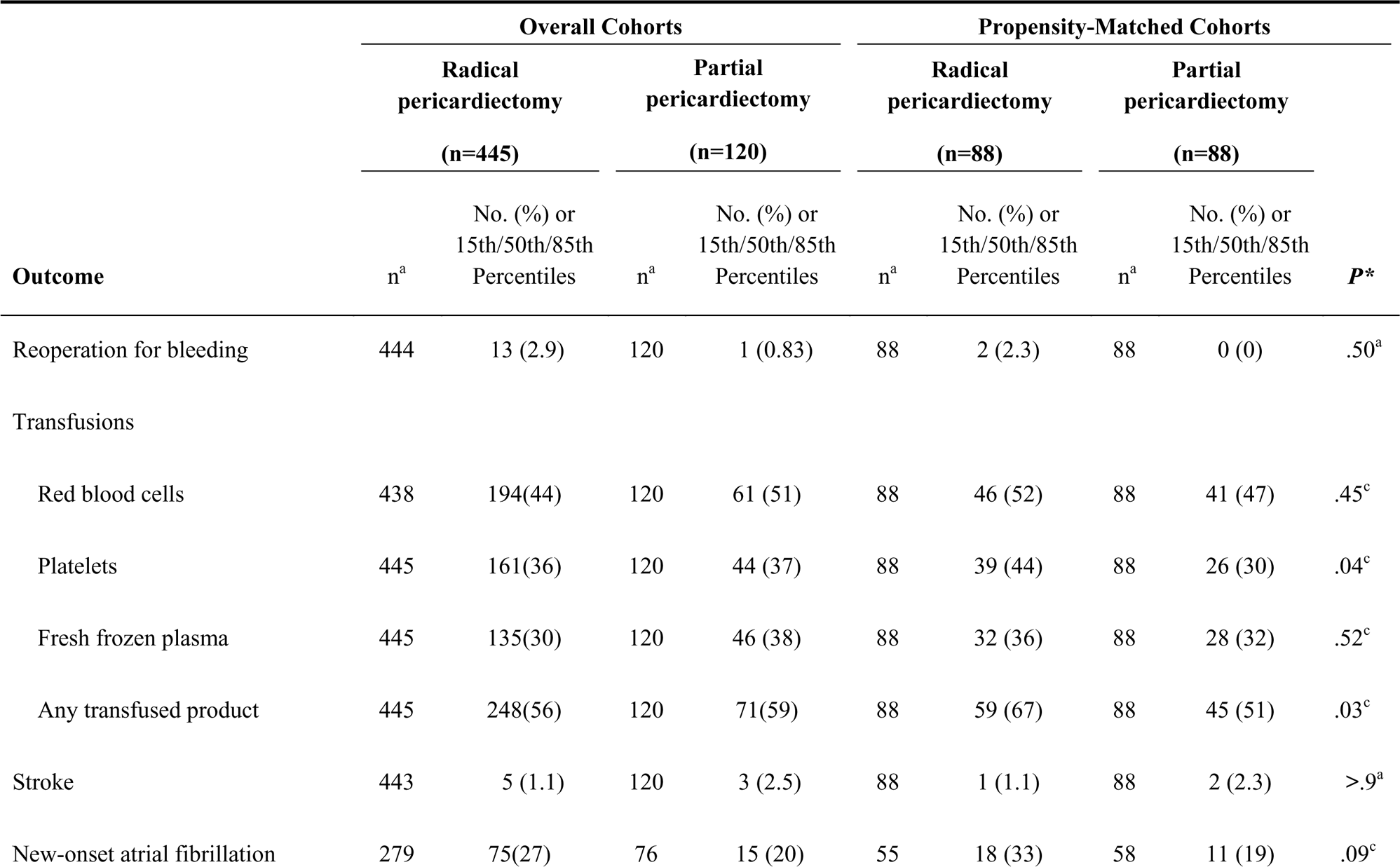
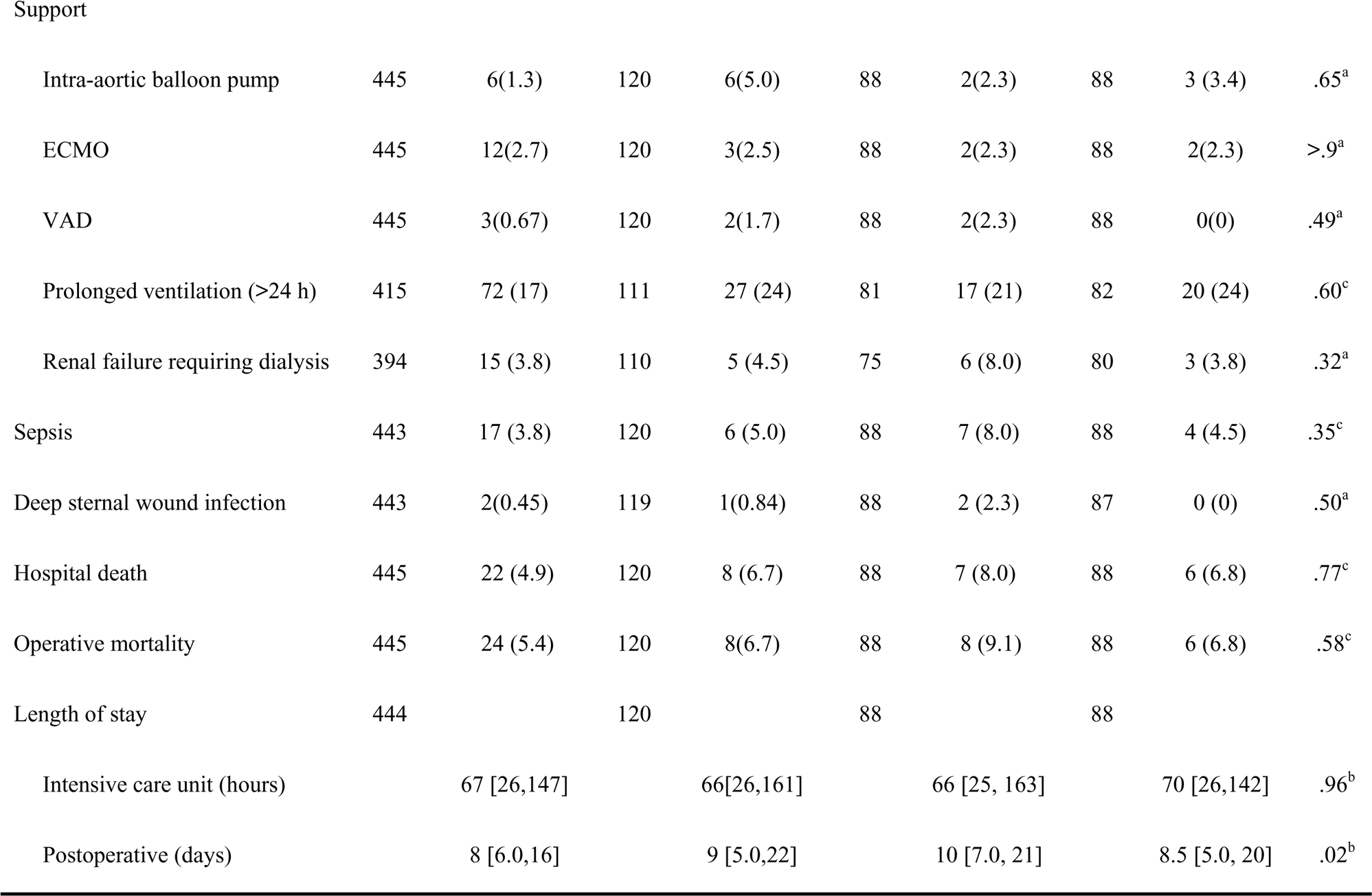
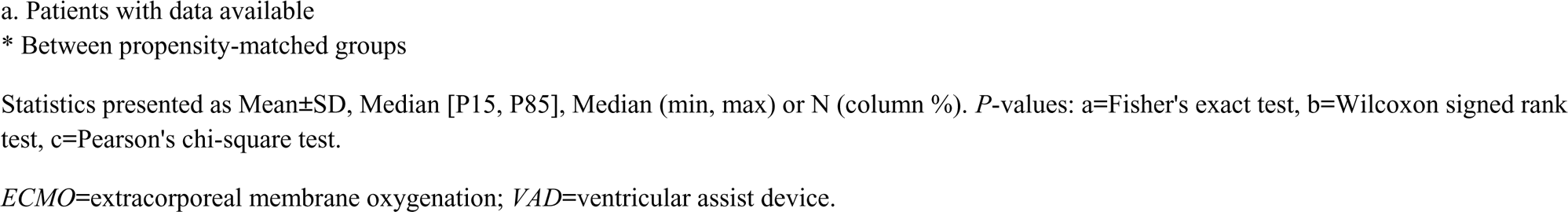
In-Hospital Outcomes.

Stroke occurred in 5 (1.1%) and postoperative atrial fibrillation in 75 (27%) (Table 4). Other complications included respiratory failure requiring prolonged intubation (n=72, 17%), new onset postoperative renal failure requiring dialysis (n=15, 3.8%), sepsis (n=17, 3.8%), and deep sternal wound infection (n=2, 0.4%).

There were 24 operative deaths (Table 4). Survival at 6 months, and 1, 5, and 10 years was 90%, 88%, 79%, and 61%, respectively (Supplemental Figure S5).

### Partial Pericardiectomy

Mean age of patients who underwent partial pericardiectomy was 61±12 years, comprised of 18 females (15%), 75 (63%) with hypertension, 38 (32%) with pharmacologically treated diabetes, 16 (14%) insulin-treated, 50 (42%) with chronic obstructive pulmonary disease, and none with chronic kidney disease.

Etiology of constrictive pericarditis was idiopathic or viral in 52 (44%), post-cardiotomy in 49 (41%), post-radiation in 14 (12%), and miscellaneous in 3 (2.5%).

Mean left ventricular ejection fraction was 55±11%. Mean preoperative cardiac index was 1.99 ±0.65 L•min^−1^•m^−2^ (Table 2).

CPB was used in 67 cases (56%) (Table 1). Intraoperatively, pericardial calcifications were observed in 48 of 100 (48%) patients (Table 3). Mean postoperative cardiac index was 3.09 ±1.13 L•min^−1^•m^−2^.

One patient (0.83%) underwent postoperative reoperation for bleeding (pericardiectomy had been performed on CPB) and 3 (2.5%) patients had a stroke (Table 4). IABP was used for postoperative support in 6 (5.0%). ECMO was used in 3 (2.5%) and LVAD in 2 patients (1.7%).

New onset atrial fibrillation occurred in 15 patients (20%) (Table 4). In 27 (24%) patients, respiratory failure required prolonged intubation. Five patients (4.5%) developed new onset postoperative renal failure requiring dialysis, and sepsis occurred in 6 (5.0%). Deep sternal wound infection occurred in 1 patient (0.84%).

There were 8 (6.7%) operative deaths. Survival at 6 months, and 1, 5, and 10 years was 87%, 84%, 64%, and 42%, respectively (Supplemental Figure S5).

### Propensity-matched Outcomes Comparison

There was a significant immediate postoperative increase in median cardiac index of 1.0 [0.10, 1.9] L•min^−1^•m^−2^, *P*<.001 in both groups (Table 3).

#### In-Hospital Morbidity

Reoperation for bleeding or tamponade occurred in 2 patients (2.3%) after radical pericardiectomy (both supported on CPB) and none after partial pericardiectomy (Table 4). Major postoperative morbidities, such as stroke, atrial fibrillation, or deep sternal wound infection, were not significantly different between radical and partial pericardiectomy groups nor was postoperative left ventricular dysfunction treated by IABP, respiratory failure requiring prolonged intubation, or renal insufficiency requiring dialysis (Table 4). Intensive care unit length of stay was similar (median: 66 vs. 70 hours; *P*=.96) although postoperative length of stay was longer (median: 10 vs. 8.5 days; *P*=.02) after radical compared to after partial pericardiectomy.

#### Early Mortality

Hospital mortality was 8.0% after radical versus 6.8% after partial pericardiectomy (*P*=.77) (Table 4). Operative mortality was 9.1% after radical versus 6.8% after partial pericardiectomy (*P*=.58).

#### Long-term Survival

Survival was 83%, 81%,71%, and 54% after radical pericardiectomy and 85%, 82%, 65%, and 41% after partial pericardiectomy at 6 months and 1, 5, and 10 years (Figure 3A). Before 5 years, survival was similar, but then diverged with better survival after radical pericardiectomy (*P*=.008) (propensity-adjusted hazard ratio 1.9 [95% CI 1.2-3.1] (Figure 3B) (Supplemental Table S2).

**Figure 3.A.**
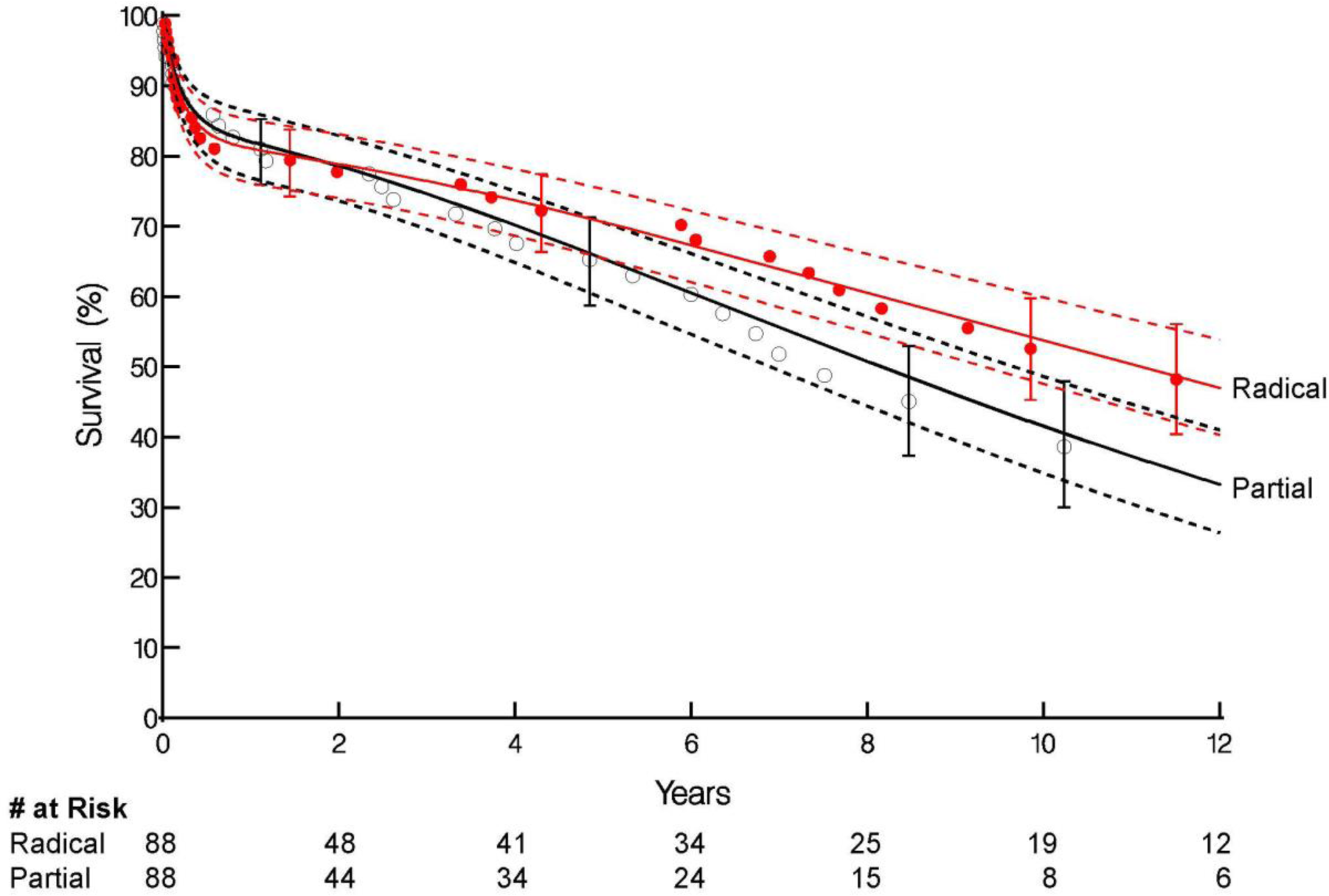
Survival After Pericardiectomy for Constriction in a Matched Cohort. Each symbol represents a Kaplan-Meier estimate of the event. Solid lines represent parametric estimates enclosed within dashed 68% confidence bands equivalent to 1 standard error. Red-Radical resection; Black-Partial resection.

**Figure 3.B.**
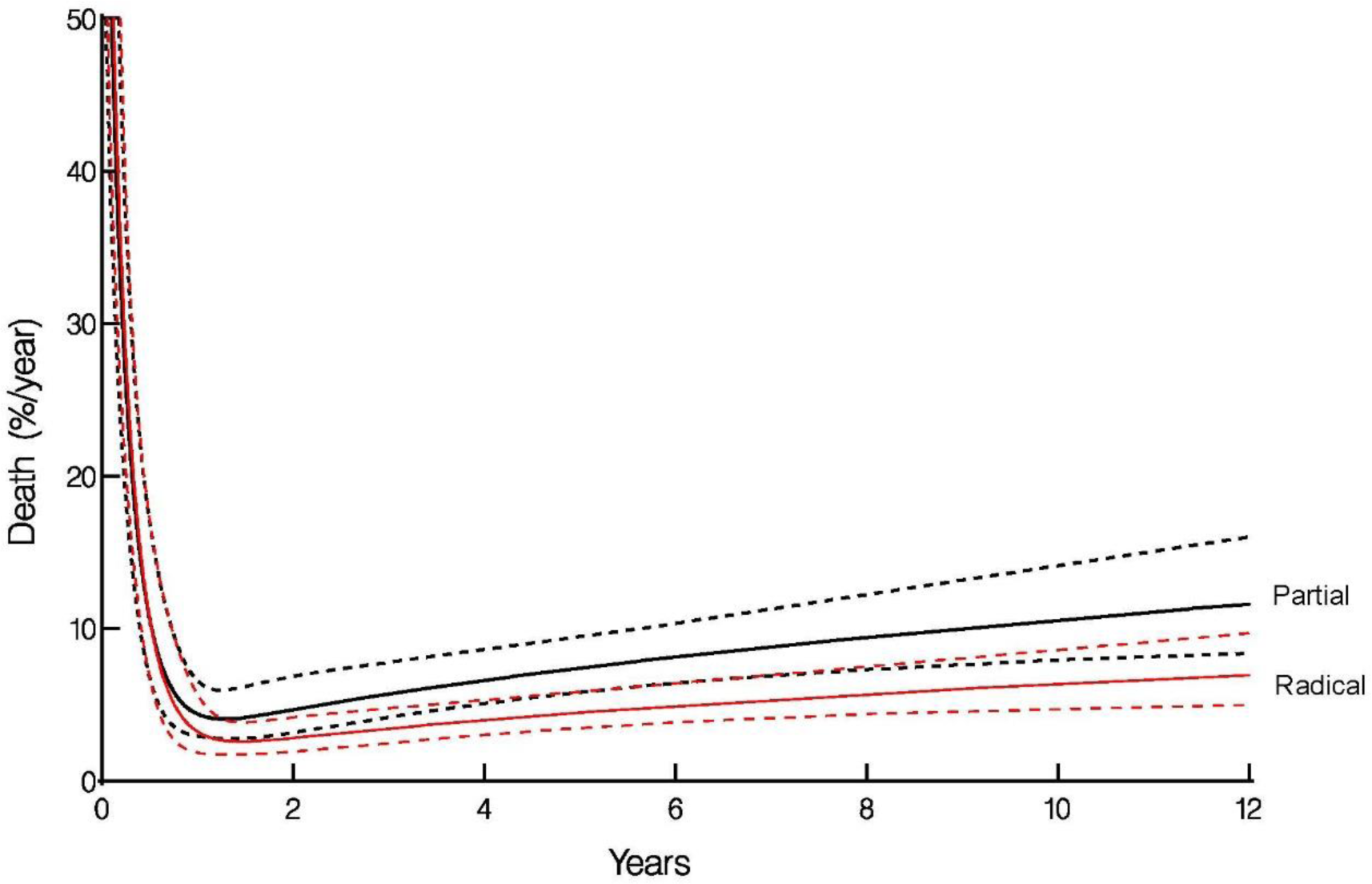
Hazard Analysis for Pericardiectomy. Each symbol represents a Kaplan-Meier estimate of the event. Solid lines represent parametric estimates enclosed within dashed 68% confidence bands equivalent to 1 standard error. Red-Radical resection; Black-Partial resection.

### Cardiopulmonary Bypass Support in Pericardiectomy

In the overall cohort, cases performed on CPB had more reoperations for bleeding (14 of 381 3.7% vs. 0% for off-pump), more transfusions (258 of 381, 68%, vs. 61/184 33%), and more postoperative atrial fibrillation (31% vs. 16%), but other outcomes were similar. Operative mortality was 23 of 381 (6.0%) with CPB and 9 of 184 (4.9%) off CPB (*P*=0.58). There was no statistically significant difference in unadjusted survival after pericardiectomy performed on versus off CPB (*P*=.49) (Supplemental Figure S6). Likewise, in the hazard model adjusting for resection type and propensity score, there were no early (*P*=.42) or late (*P*=.65) differences in survival with or without use of CPB (Table 5).

**Table 5.**
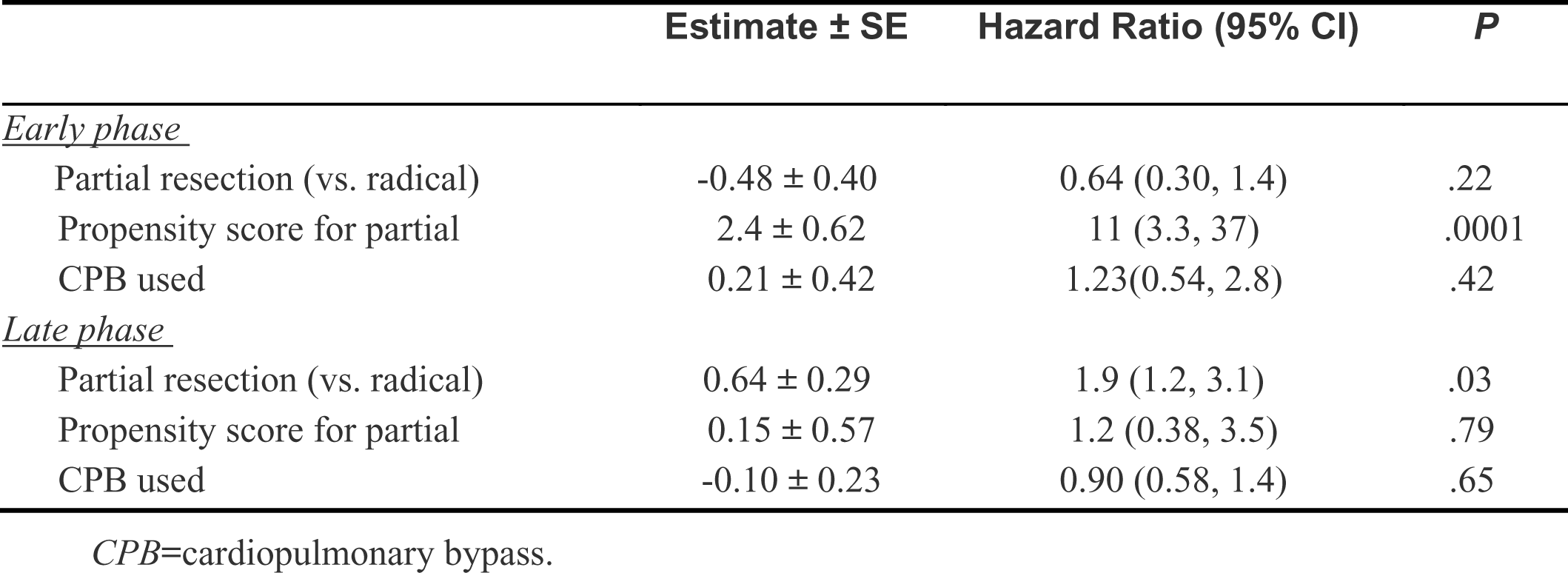
Hazard Model for Death After Propensity Adjustment, Incorporating CPB Use (n=565).

## DISCUSSION

### Principal Findings

Both radical and partial pericardiectomy for constrictive pericarditis provide similar short-term hemodynamic relief demonstrated by significant postoperative cardiac index increase in both groups. Performing radical pericardiectomy in patients provides better long-term survival than partial pericardiectomy, without compromising early outcomes such as perioperative morbidity or mortality. CPB does not substantially increase the morbidity of the procedure, with exception of greater number of transfusions and reoperations for bleeding or tamponade.

### Cleveland Clinic Historical Perspective

In the 1990s, Lytle and colleagues performed surgical pericardiectomy at Cleveland Clinic.^2^ At that time, it was considered surgical dogma to avoid CPB due to increased risk of postoperative bleeding. In addition to median sternotomy, bilateral thoracotomies and clamshell incisions were used to improve exposure and different types of hemodynamic support were used during heart manipulation. In the 2000s, Smedira and colleagues introduced early CPB initiation. This provided hemodynamic stability during surgery, thereby enabling more extensive radical/circumferential pericardial resection that included pericardium around the IVC and pulmonary veins. This evolved into the current radical pericardiectomy as described in Methods.^1^

### Mortality and Morbidity

Early mortality after pericardiectomy is related to era of surgery, and particularly to etiology of constrictive pericarditis. In a study documenting more than 80 years of experience following pericardiectomy outcomes at Mayo Clinic, Murashita and colleagues reported a decline in mortality from 13% in historical patients to 5% since 1990.^20^ This trend is consistent with the findings of the present study, which show an overall mortality of 5.6%, despite a more aggressive approach to operating on patients with prior cardiac surgery requiring concomitant valve and coronary operations.

In our study, reoperation for bleeding was low. Concern for bleeding has been reported as a reason to limit resection and use of CPB.^21,22^ Renal failure of 5.5% may reflect presence of longstanding right heart failure in this population along with subclinical cirrhosis and deserves further study.

A unique and devastating complication of pericardiectomy is acute over-distension of the right ventricle following pericardium removal. When this occurs, it can lead to rapid progression of cardiogenic shock and death or need for mechanical circulatory support.^23^ With routine use of fluid restriction and low-dose inotropes to limit ventricular volume after resection, 2.6% of patients required IABP and 5.1% were supported temporarily with ECMO.

### Hemodynamics and Outcomes

Clinical success after pericardiectomy for constrictive pericarditis depends on a complex interplay of factors, including presence of coexistent valve, coronary, or myocardial disease. However, successful release of diastolic restraint can result in immediate improvement in cardiac index. Patients undergoing radical pericardiectomy compared with those undergoing partial pericardiectomy had lower preoperative cardiac index, which may indicate that radical resection was employed in patients with more advanced constriction. At the same time, they had better survival after 5 years. We hypothesize that the observed survival benefit might be attributed to recurrent pericarditis on the residual pericardium with possible re-constriction. Low postoperative cardiac index was strongly correlated with worse survival (Supplemental Figure S6) and may indicate the presence of restrictive cardiomyopathy, which may be misdiagnosed as constriction, or other underlying conditions masked by constrictive pericarditis.

The ratio of right atrial pressure and pulmonary capillary wedge pressure might be a useful tool in the assessment of myocardial involvement in constriction and might predict postoperative outcomes.^24^ Further analysis could enhance our ability to predict the patients who are less likely to benefit from pericardiectomy, as in the case of post-radiation constriction.

The major determinant of long-term survival after pericardiectomy for constrictive pericarditis was etiology, as observed in the previous experience by Bertog and colleagues^2^ and in other large series.^21^ Patients with idiopathic constriction, even in the setting of advanced calcification and unfavorable preoperative hemodynamics, have excellent short- and long-term survival. Those with post-surgical constriction had worse early and late outcomes, which may be indicative of multifactorial, mixed pericardial/myocardial disease and not just pericardial constriction.^7^ Patients with radiation-induced constrictive pericarditis had the worst outcomes, not surprising given coexistence of ventricular fibrosis, valvular and coronary disease, and radiation-induced pulmonary fibrosis in many of them. These findings support need for a different approach to timing and operative planning in different patient populations. Patients with idiopathic pericarditis may benefit from aggressive early surgery given the excellent short- and long-term survival, whereas patients who have had radiation treatment merit extreme caution.

### Completeness of Resection

European Society of Cardiology 2015 Guidelines recommend a tailored approach to pericardial resection, stating debate between radical and partial resection is now unnecessary.^10^ However, a suggestion is made that resection may be limited in the presence of severe calcification. Advocates of limited resection suggest partial pericardiectomy results in acceptable outcomes.^25^ Although partial resection can result in symptomatic improvement, some patients develop recurrent symptoms from continued constriction of the posterior pericardium and require redo pericardiectomy. Cho and colleagues report a series of radical pericardiectomies for recurrent constriction. Partial pericardiectomy performed as initial surgery was the most common reason for recurrence.^7^ Murashita and colleagues recommend as complete a resection as possible, leaving a strip of pericardium preserved along the phrenic nerve.^20^

In concordance, radical versus partial pericardiectomy in matched cohorts did not show increased hospital mortality or worse survival in this study. In fact, radical resection was associated with better late long-term survival. Whether radical resection results in better freedom from recurrent symptoms requires further study. In the interim, these results should inspire confidence that radical resection does not increase the risk of the procedure. This means there is no reason not to maximize removal of pericardium, including scar and calcific tissue, as long as deemed safe — even if this requires CPB.

### Role of Cardiopulmonary Bypass

Use of CPB remains controversial even among surgeons who advocate radical pericardial resection. In multiple reports, the use of CPB has been associated with worse outcomes, even in the hands of experienced surgeons.^9,26^ Assessing the incremental risk of CPB in pericardiectomy is challenging because, at least historically, it has often been employed as a rescue strategy in the case of catastrophic bleeding to mitigate hemodynamic instability during resection, or to support concomitant procedures that may increase risk or reflect underlying cardiac disease. If CPB is used only in more complex cases, it would be expected to be a marker for increased severity and risk.

The range of opinions for using CPB seems to be: 1) avoid it completely; 2) use it when necessary to repair bleeding or perform concomitant procedures; or 3) use it routinely to facilitate radical resection. Evolution in our practice started as avoidance of CPB to more liberal use and now routine use of CPB as almost a condition for radical pericardiectomy, which aids in avoiding accidental cardiac injury, facilitates maintaining end-organ perfusion during dissection of the lateral left ventricle and diaphragmatic surface, and permits a more controlled resection. Surgeons who are less comfortable navigating the often-difficult lateral and posterior pericardium dissection may find CPB beneficial as an adjunct to allow for more complete resection. Use of bipolar, irrigated hemostatic sealer has made a substantial difference in the control of bleeding after CPB and in these more extensive radical dissections, minimizing the bleeding complications.

In our study, when all patients were considered, there was an increased and statistically significant risk for reoperation for bleeding associated with CPB. However, after matching there was no statistically significant difference in this outcome. This could potentially be explained by the low bleeding rates we found. As CPB use becomes more prevalent, the low rates of reoperation for bleeding raise the question about the clinical significance of this finding.

### Limitations

This clinical study reviews a heterogeneous patient population. The gradual evolution of approaches and pericardial surgical techniques makes it difficult to determine whether it was the surgeon’s intent to use a particular surgical strategy, such as CPB or radical resection, or whether this was guided by intraoperative findings. Although the gradual evolution of technique makes adequate comparison difficult, it is important to note that neither surgical approach was associated with a difference in early mortality. One important limitation is lack of long-term functional status data, which is especially relevant for this disease that can produce symptoms out of proportion to physical findings or imaging.

### Conclusions

The current surgical approach to pericardiectomy at our institution reflects a gradual, parallel evolution towards more liberal use of CPB and more aggressive pericardial resection. Regardless of the approach and completeness of resection, pericardiectomy for idiopathic constrictive pericarditis is associated with low mortality and morbidity, and results in excellent long-term survival. Among matched patients, better long-term survival with radical versus partial pericardiectomy suggests that it should be the standard of care in the surgical treatment of constrictive pericarditis. Similarly, use of CPB is low-risk and beneficial in these difficult operations.

## Data Availability

Clinical data were extracted prospectively for quality assurance. Cleveland Clinic's Institutional Review Board approved use of these data for research, with patient consent waived (IRB #5001; approved 2/26/2021). Data can be provided for quality assurance and review with appropriate protocol, processes and mutual agreement.

## ABBREVIATIONS

COPD: chronic obstructive pulmonary disease
CPB: cardiopulmonary bypass
ECMO: extracorporeal membrane oxygenation
IABP: intra-aortic balloon pump
IVC: inferior vena cava
LVAD: left ventricular assist device
SVC: superior vena cava

## ACKNOWLEDGEMENTS

The authors thank Ingrid Schaefer Sprague for her editorial assistance.

